# Mild Behavioral Impairment and Cortical Thinning: Biomarkers of Early Neurodegeneration

**DOI:** 10.1101/2024.12.19.24319306

**Authors:** Yi Jin Leow, Seyed Ehsan Saffari, Ashwati Vipin, Pricilia Tanoto, Rasyiqah Binte Shaik Mohamed Salim, Bocheng Qiu, Zahinoor Ismail, Nagaendran Kandiah

## Abstract

**Background:** Mild Behavioral Impairment(MBI) is increasingly recognized as an early phenotypic marker of neurodegeneration, with neuropsychiatric symptoms(NPS) frequently emerging prior to cognitive deficits. While structural neuroimaging studies suggest a link between cortical thinning and NPS, the link between MBI and cortical morphology remains underexplored in diverse community-based cohorts. This study investigated whether early behavioral alterations, assessed via the Mild Behavioral Impairment Checklist(MBI-C), correlate with region-specific cortical thinning in a Southeast Asian cohort.

**Methods:** We conducted a cross-sectional analysis of 969 participants (mean age 61.99±10.19years;39.6% male;87.2%Chinese) enrolled in the Biomarkers and Cognition Study in Singapore(BIOCIS), ranging from cognitively normal, subjective cognitive decline(SCD), to mild cognitive impairment(MCI). MBI was quantified using the self-reported MBI-C. T1-weighted MRI scans were processed with FreeSurfer to measure cortical thickness. Associations between MBI-C scores(total/subdomains) and cortical thinning were examined.

**Results:** Higher MBI-C Belief scores were significantly associated with cortical thinning in the right hemisphere(β=−0.0177;95%CI:−0.0342to−0.0012;P=0.035). Region-specific analyses showed significant thinning in posterior banks of the superior temporal sulcus, fusiform gyrus, superior temporal gyrus, temporal pole, and transverse temporal gyrus, which remained significant after false discovery rate correction(FDR P=0.042–0.045). Additional thinning was noted in right postcentral and supramarginal gyri and right insula(FDR P≤0.039).

**Conclusions:** Elevated MBI, particularly abnormal beliefs, is linked to cortical thinning in regions subserving memory, sensory integration, and emotional regulation, especially within the right hemisphere. These findings highlight the potential of MBI-C as an early neurodegenerative marker and underscore the need for longitudinal studies to clarify temporal dynamics and mechanisms underlying behavioral symptoms and neurodegenerative processes.

---

The brain and its behavior are fundamental to our humanity. Research in these areas deepens our knowledge of what it means to be human and aids in development of treatments for neurological and psychiatric conditions. Recent findings suggest that neuropsychiatric symptoms (NPS) and, its subset Mild Behavioral Impairment (MBI) not only presage cognitive deterioration but also reflect fundamental alterations in brain morphology (1–3). Emerging evidence reveals that both behavioral disturbances and structural brain changes occur well before measurable cognitive deficits emerge, thereby accurately forecasting the onset of Alzheimer’s disease (AD) and related dementias (4–6). Consequently, integrative, multimodal approaches that combine behavioral assessments with neuroimaging biomarkers are critical for the early detection and diagnosis of AD and other neurodegenerative disorders.

In clinical practice, neuropsychological assessments remain the cornerstone for evaluating cognitive performance and diagnosing impairment. However, they are increasingly complemented by neuropsychiatric evaluations and neuroimaging studies, which capture behavioral and structural changes that precede cognitive symptoms. NPS refers to a broad range of non-cognitive, behavioral, or psychiatric symptoms that frequently precede or accompany cognitive decline. Baseline NPS, such as depression, anxiety, and agitation, have been identified as significant predictors of incident mild cognitive impairment (MCI) and dementia in older adults (7–9). Furthermore, over 59% of individuals with MCI or AD exhibit NPS prior to diagnosis, with depression (24%) and irritability (21%) being among the most prevalent early symptoms(10). A review of 61 studies revealed that depression, present in 45.9% of cases, is a significant predictor of the transition from preclinical to prodromal stages of dementia (4). Further, Ferreira et al. (2023) demonstrated that NPS can forecast the etiology, severity, and progression of AD, reinforcing their role as early indicators of cognitive impairment.

Within the spectrum of NPS, MBI has been identified as a distinct syndrome that captures early behavioral changes predictive of cognitive decline and dementia (11,12). Notably, MBI has been incorporated into the National Institute on Aging and Alzheimer’s Association (NIA-AA) AD research framework as a potential preclinical indicator of underlying neuropathology (13). Current literature indicates that MBI is a robust and independent predictor of dementia, irrespective of baseline cognitive function. (14–16). The Mild Behavioral Impairment Checklist (MBI-C) developed by (17), systematically categorizes MBI into five domains—decreased motivation, affective dysregulation, impulse dyscontrol, social inappropriateness, and abnormal perception or false beliefs—each of which is linked to subsequent cognitive decline (18). A four-year longitudinal study demonstrated that integrating the MBI-C into clinical assessments significantly enhances the predictive accuracy for dementia (19). Notably, the MBI-C is sensitive to the earliest detectable behavioral changes in dementia-free individuals, making it a powerful tool for identifying prodromal neurodegenerative changes (20,21).

Beyond behavioral assessments, neuroimaging biomarkers play a crucial role in detecting and monitoring early neurodegeneration. Cortical thickness measurement via magnetic resonance imaging (MRI) has emerged as a particularly sensitive and reproducible marker of neuronal integrity, capturing subtle regional variations associated with both normal aging and early disease-related changes (22,23). Recent studies have validated cortical thickness as a reliable biomarker, demonstrating high sensitivity and specificity in detecting early neurodegenerative change and memory changes (24,25). This method consistently reveals progressive cortical thinning in individuals with cognitive impairment and dementia, particularly in regions critical for memory.

Longitudinal analyses have further established that thinning in the medial temporal, lateral temporal, parietal, and frontal cortices reliably predicts cognitive decline, with greater atrophy corresponding to accelerated deterioration in memory, executive function, and global cognition (26–28). Notably, cortical thinning observed in asymptomatic, amyloid-positive individuals heralds early preclinical Alzheimer’s disease pathology, underscoring its potential as a non-invasive early biomarker. Furthermore, significant thinning in regions susceptible to amyloid deposition, such as the precuneus and posterior cingulate cortex, is evident even during presymptomatic stages, reflecting early neurodegenerative processes (29,30). In addition, reductions in cortical thickness have been shown to correlate with amyloid burden and predict future cognitive decline, thereby identifying individuals at elevated risk for progression to AD (31). Given its reproducibility, sensitivity, and predictive value, cortical thickness measurement has emerged as a critical imaging biomarker in the assessment of neurodegenerative diseases.

Recent research underscores the link between NPS, MBI and and cortical thinning. Gill et al. (32) found that impulse dyscontrol in MBI is linked to thinning in the orbitofrontal cortex, anterior cingulate cortex (ACC) and insula, regions associated with poorer cognitive outcomes and dementia progression. Similarly, reduced cortical thickness and increased white matter hyperintensities have been observed in NPS across neurodegenerative and cerebrovascular diseases (33). In the study by Donovan et al, cortical thinning in regions like the medial frontal cortex, ACC, and inferior parietal lobule predicts worsening apathy and hallucinations along the AD spectrum (34). Apathy has also been associated with lower inferior temporal thickness in both MCI patients and cognitively normal elderly individuals, suggesting its potential as an early marker of degeneration (35).

Findings from Asian cohorts further reinforce these associations. In Japan, Imai et al. (36) observed that emotional dysregulation in individuals with MBI was linked to thinning in the right supramarginal gyrus, suggesting early neurodegenerative changes. Likewise, in Korea, Yoon et al. (3) found that higher MBI-C scores were associated with progression from amnestic MCI to Alzheimer’s disease, with cortical thinning observed in temporal, parietal, and frontal regions. In addition, a Korean multi-center study by Kim et al. (37) reported that frontal behavioral impairments were correlated with more severe thinning and faster cognitive decline, particularly in executive function and memory Despite these advancements, prior studies on cortical thickness and NPS have predominantly relied on clinical samples (n = 100–500) using the Neuropsychiatric Inventory (NPI) or NPI-Q, which primarily assess dementia-related behaviors over short reference periods (2–4 weeks) and are designed to capture later-stage symptoms (e.g., biting, wandering). In contrast, the MBI-C evaluates symptoms over a six-month period, allowing transient states (e.g., stress or medication effects) to subside and thus providing a more accurate measure of sustained impairments (17). This extended timeframe enhances the early identification of NPS, thereby improving diagnostic and prognostic accuracy in at-risk populations.

To our knowledge, this is the first study to leverage the MBI-C in assessing cortical thickness in a community-dwelling Southeast Asian cohort. This study aims to determine whether early behavioral changes captured by the MBI-C are associated with region-specific cortical thinning in a multi-ethnic cohort, thereby providing novel insights into the structural correlates of early behavioral changes and neurodegenerative processes. Given the growing evidence that behavioral symptoms serve as prodromal markers of cognitive decline, we hypothesize that early behavioral changes, as quantified by the MBI-C, may function as biomarkers of neurodegeneration.

## Methods

### Biomarkers and Cognition Study, Singapore

Cross-sectional data were drawn from the Biomarkers and Cognition Study, Singapore (BIOCIS), which recruits community-dwelling participants in Singapore. To investigate underlying pathology in individuals with and without minimal symptoms, BIOCIS includes both cognitively impaired and unimpaired participants. Following Petersen’s (Petersen, 2004) criteria and the National Institute on Aging–Alzheimer’s Association (NIA-AA) guidelines, based on neuropsychological assessments and self-reported memory issues, participants are classified as Cognitively Normal (CN), Subjective Cognitive Decline (SCD), or MCI. Inclusion criteria are as follows: (1) age 30–95 years; (2) literacy in English or Mandarin; and (3) intact mental capacity. Exclusion criteria include: (1) any serious neurological, psychiatric, or systemic disease; (2) history of psychotic disorder or major depressive disorder; and (3) alcoholism or drug dependency/abuse within the past two years. Further details on methodology can be found in the publoished protocol paper (Leow et al., 2024). For this study, only participants with baseline data for all covariates of interest (e.g., years of education, cognitive diagnosis, MBI-C scores, cortical thickness data; see Figure 1) were included.

**FIGURE 1.**
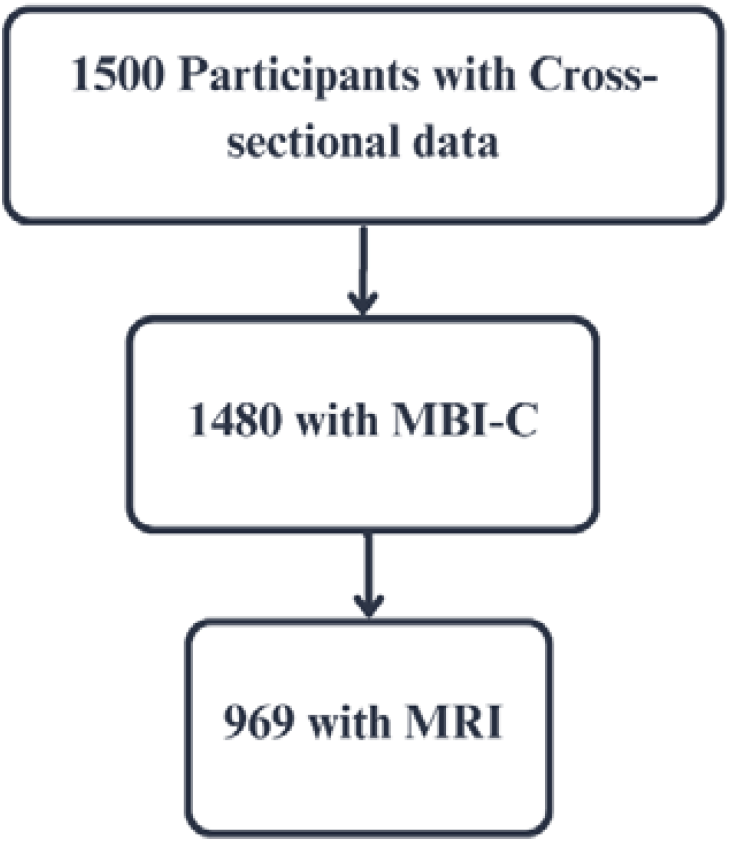
Flowchart illustrating sample populations were obtained from the BIOCIS Cohort. BIOCIS = Biomarkers and Cognition Study, Singapore, MBI-C = Mild Behavioral Impairment-Checklist, MRI = Magnetic Resonance Imaging.

### Mild Behavioral Impairment-Checklist (MBI-C)

The MBI-C (Ismail et al., 2017), available at MBItest.org, consists of 34 items across five domains: decreased motivation (6 items), affective dysregulation (6 items), impulse dyscontrol (12 items), social inappropriateness (5 items), and abnormal perception (5 items). Each item is scored from 0 (absent) to 3 (severe), yielding a total score of 0–102. Participants self-reported symptoms lasting at least six months, reflecting changes from prior behavior.

### Neuroimaging Acquisition and Processing

All participants underwent whole□brain MRI scans using a 3T Siemens Prisma Fit scanner (Siemens Healthineers, Erlangen, Germany). A T1-weighted MPRAGE sequence was used with the following parameters: repetition time=2000 ms, echo time=2.26 ms, inversion time=800 ms, flip angle=8°, matrix size=256×256, and voxel size=1.0×1.0×1.0 mm^3^. Scans were processed with FreeSurfer (v7.2.0) using the “ recon-all” pipeline, which included motion correction, removal of non-brain tissue, automated Talairach transformation, intensity correction, volumetric segmentation, and cortical surface reconstruction and parcellation (Desikan et al., 2006; Fischl et al., 2002; Fischl, Salat, et al., 2004; Fischl, Van Der Kouwe, et al., 2004). The FreeSurfer cortical thickness estimations were obtained using FreeSurfer default automated settings. As per the Desikan-Killiany atlas (38) the cortical thickness from 34 brain regions were extracted (separately for the left and right hemispheres) into 6 regions: Frontal Lobe (Caudal Middle Frontal, Lateral Orbitofrontal, Medial Orbitofrontal, Pars Opercularis, Pars Orbitalis, Pars Triangularis, Precentral, Rostral Middle Frontal, Superior Frontal, Frontal Pole); Parietal Lobe (Inferior Parietal, Paracentral, Postcentral, Precuneus, Superior Parietal, Supramarginal); Temporal Lobe (Posterior Banks of Superior Temporal Sulcus, Entorhinal, Fusiform, Inferior Temporal, Middle Temporal, Parahippocampal, Superior Temporal, Temporal Pole, Transverse Temporal); Occipital Lobe (Cuneus, Lateral Occipital, Lingual, Pericalcarine); Cingulate Cortex (Caudal Anterior Cingulate, Isthmus Cingulate, Posterior Cingulate, Rostral anterior Cingulate) and Other Regions (Insula). Mean cortical thickness for each of the regions mentioned were calculated separately for the right and left hemispheres.

### Statistical Analyses

Continuous variables were summarized as mean and standard deviation (SD), while categorical variables were expressed as frequency and percentage. All continuous variables were screened for normality using the Shapiro-Wilk test, and those demonstrating skewness were log-transformed (with a constant of 1 added for values <1) to ensure appropriate normalization. Group comparisons of continuous variables between cognitive groups were performed using independent sample t-tests or the Mann-Whitney U test, depending on these normality assumptions. Chi-square tests (or Fisher’s exact test for low expected cell counts) were used to compare categorical variables. To examine the relationship between cortical thickness and MBI-C scores, multivariable linear regression models were employed, adjusting for potential confounders such as age, education, gender, apolipoprotein E4 (APOE ε4) and global cognitive function as measured by the Montreal Cognitive Assessment (MoCA).

Previous research suggests that reductions in cortical thickness may correlate with both cognitive decline and behavioral disturbances. However, it remains unclear whether emerging behavioral changes directly reflect underlying structural alterations or are secondary to cognitive deficits. To more precisely isolate the contribution of cortical structure, MoCA scores were included as a covariate in all relevant analyses—consistent with previously published protocols (3,36) —thus disentangling structural effects from general cognitive impairment.

Regression analyses reported standardized Beta coefficients (β) to facilitate comparability of effect sizes and enable nuanced interpretation of results. Additionally, 95% confidence intervals (CIs) and p-values were provided to quantify the precision and statistical significance of associations. All analyses were two-sided, with the significance threshold set at p < 0.05 unless otherwise specified. The BIOCIS study was adequately powered for its primary endpoints, allowing for exploration of relationships between cortical thickness and MBI-C scores as secondary endpoints. To control for multiple comparisons and avoid inflation of type I error, False Discovery Rate (FDR) correction was applied in Table 3 to detailed associations between cortical thinning in specific brain subregions and MBI-C Belief Scores, with adjusted p-values reported. All analyses were conducted using IBM SPSS Statistics for Windows, Version 29.0 (IBM Corp., Armonk, NY).

## Results

### Participant demographics

Data from 969 participants (mean age 61.99 years ± 10.19, 39.6% males, 87.2% Chinese) were analyzed. 34.3% was classified as CN, 26.4% as SCD, and 39.3% as MCI. Table 1 presents descriptives of demographics, cognitive, and cortical thickness profiles of the BIOCIS cohort.

**Table 1.** Descriptives of Demographics, Global Cognition, Behavioral and Cortical Thickness Profiles.

|  | CN<br>n=332 | SCD<br>n=256 | MCI<br>n=381 | Overall<br>n=969 | Overall<br>*p* □ |
| --- | --- | --- | --- | --- | --- |
| **Age†** | 54.88±10.31 | 56.93±9.27 | 62.94±10.25 | 58.59±10.64 | **<.001** |
| **Sex (Male) □** | 122 (37.1%) | 101(39.45%) | 161 (42.3%) | 384 (39.8%) | 0.37 |
| **Education†** | 15.74±3.2 | 15.07±3.03 | 13.93±3.88 | 14.85±3.53 | **<.001** |
| **MoCA†** | 27.03±2.18 | 26.24±2.41 | 25.01±2.81 | 26.03±2.65 | **<.001** |
| **MBI-C Total** | 1.38 ± 2.912 | 3.93 ± 7.088 | 2.59±4.763 | 2.53 ± 5.102 | **<.001** |
| **MBI-C Interest** | 0.29 ± 0.897 | 0.93 ± 1.985 | 0.47±1.193 | 0.53 ± 1.392 | **<.001** |
| **MBI-C Mood** | 0.41 ± 1.193 | 1.12 ± 2.011 | 0.75±1.523 | 0.73 ± 1.594 | **<.001** |
| **MBI-C Control** | 0.58 ± 1.261 | 1.44 ± 2.748 | 1.08±2.418 | 1.00 ± 2.223 | **<.001** |
| **MBI-C Social** | 0.08 ± 0.338 | 0.22 ± 0.885 | 0.17 ± 0.565 | 0.15 ± 0.612 | **0.026** |
| **MBI-C Beliefs** | 0.02 ± 0.189 | 0.22 ± 0.907 | 0.11 ± 0.540 | 0.11 ± 0.591 | **<.001** |
| **Cortical Thickness (Left hemisphere)** |  |  |  |  |  |
| **Temporal†** | 2.3374±0.1 | 2.326±0.1 | 2.3023±0.11 | 2.3206±0.1 | **<.001** |
| **Cingulate Cortex†** | 2.4478±0.11 | 2.4274±0.1 | 2.4226±0.11 | 2.4325±0.11 | 0.005 |
| **Frontal†** | 2.4835±0.09 | 2.4794±0.08 | 2.4804±0.09 | 2.4812±0.09 | 0.835 |
| **Occipital†** | 1.8262±0.08 | 1.819±0.09 | 1.8081±0.09 | 1.8172±0.09 | 0.017 |
| **Parietal†** | 2.2962±0.1 | 2.2805±0.09 | 2.2583±0.11 | 2.2772±0.1 | **<.001** |
| **Other Regions - Insula†** | 2.9142±0.13 | 2.8846±0.12 | 2.8728±0.14 | 2.8901±0.13 | **<.001** |
| **Mean of Hemisphere†** | 2.4015±0.08 | 2.3894±0.07 | 2.3745±0.09 | 2.3877±0.08 | **<.001** |
| **Cortical Thickness (Right hemisphere)** |  |  |  |  |  |
| **Temporal†** | 2.3691±0.1 | 2.3552±0.1 | 2.3298±0.11 | 2.35±0.11 | **<.001** |
| **Cingulate Cortex†** | 2.3971±0.12 | 2.3862±0.11 | 2.3781±0.12 | 2.3867±0.12 | 0.088 |
| **Frontal†** | 2.4673±0.09 | 2.4621±0.07 | 2.4734±0.09 | 2.4683±0.09 | 0.272 |
| **Occipital†** | 1.8556±0.08 | 1.8515±0.09 | 1.8383±0.09 | 1.8477±0.09 | 0.019 |
| **Parietal†** | 2.3071±0.1 | 2.2883±0.09 | 2.2661±0.12 | 2.286±0.11 | **<.001** |
| **Other Regions - Insula†** | 2.9217±0.14 | 2.9047±0.13 | 2.885±0.15 | 2.9028±0.14 | 0.002 |
| **Hemisphere†** | 2.3961±0.08 | 2.384±0.07 | 2.3702±0.09 | 2.3827±0.08 | **<.001** |

*Note.* P values significant at $\leq 0.05$ are bolded, † = mean ± Standard Deviation, □ = Frequency (percentages), □ = non-adjusted. Cortical data is presented in millimetres.
Abbreviations: APOE ε4 = apolipoprotein E, CN = Cognitively Normal , MBI-C = Mild Behavioral Impairment-Checklist, MCI = Mild Cognitive Impairment, MoCA = The Montreal Cognitive Assessment, SCD = Subjective Cognitive Decline.

Significant group differences were observed for age (p < .001), education (p < .001), and MoCA (p < .001). All MBI-C domains differed significantly: Total (p < .001), Interest (p < .001), Mood (p < .001), Control (p < .001), Social (p = .026), and Beliefs (p < .001). For left hemisphere cortical thickness, significant differences emerged in the temporal (p < .001), cingulate (p = .005), occipital (p = .017), parietal (p < .001), insula (p < .001), and overall mean (p < .001) measures, but not in the frontal region (p = .835). In the right hemisphere, significant differences were found in the temporal (p < .001), occipital (p = .019), parietal (p < .001), insula (p = .002), and overall mean (p < .001) measures, whereas the cingulate (p = .088) and frontal regions (p = .272) did not differ significantly.

### Cortical thickness and Mild Behavioral Impairment

Significant associations were found between cortical thinning and elevated MBI-C Belief scores, particularly in the right hemisphere. In the left hemisphere, none of the associations with MBI-C domains reached statistical significance. In the right hemisphere, cortical thinning was significantly associated with higher MBI-C Belief scores (β = -0.01768, 95% CI: -0.03416, -0.0012, p = 0.035). This indicates that a one standard deviation (SD) decrease in overall cortical thickness in the right hemisphere corresponds to a 0.01768-point increase in MBI-C Belief scores. Results are summarized in Table 2, with detailed findings on MBI-C domains and specific brain regions provided in Supplementary Table 1.

**Table 2.** Relationship Between Hemispheric Cortical Thickness and MBI in the BIOCIS Cohort.

|  | Left hemisphere means |  |  | Right hemisphere means |  |  |
| --- | --- | --- | --- | --- | --- | --- |
|  | B | 95% CI | p | B | 95% CI | p |
| **MBI-C Total** | 0.01016 | -0.05208, 0.07232 | 0.749 | 0.0044 | -0.05696, 0.06568 | 0.888 |
| **MBI-C Interest** | -0.01472 | -0.05024, 0.0208 | 0.416 | -0.02144 | -0.05648, 0.01352 | 0.229 |
| **MBI-C Mood** | 0.01392 | -0.02528, 0.05304 | 0.486 | 0.01368 | -0.02488, 0.05232 | 0.487 |
| **MBI-C Control** | 0.01328 | -0.03088, 0.05736 | 0.556 | 0.00408 | -0.03944, 0.04752 | 0.855 |
| **MBI-C Social** | -0.00864 | -0.02776, 0.0104 | 0.373 | -0.00864 | -0.02744, 0.01016 | 0.365 |
| **MBI-C Beliefs** | -0.012 | -0.02872, 0.00472 | 0.159 | -0.01768 | -0.03416, -0.0012 | **0.035** |

*Note.* Models were adjusted for age, education, APOE ε4, and global cognition scores. Significant p-values appear bold.
Abbreviations: APOE ε4 = apolipoprotein E4, BIOCIS = Biomarkers and Cognition Study, Singapore, MBI-C = Mild Behavioral Impairment-Checklist

Further analysis of the significant MBI-C subdomain identified in Table 2 revealed additional associations between specific subregions and MBI-C-Belief scores. In the left hemisphere, thinning in the entorhinal cortex was associated with higher MBI-C Belief scores (β = -0.017, 95% CI: -0.032, -0.001, p = 0.034), although this did not remain significant post-FDR correction (FDR p = 0.153). Similarly, thinning in the fusiform gyrus (β = -0.020, 95% CI: -0.036, -0.004, p = 0.014) lost significance after correction (FDR p = 0.126).

In the right hemisphere, multiple regions exhibited significant associations with MBI-C Belief scores. These included the posterior banks of the superior temporal sulcus (β = -0.020, 95% CI: -0.037, -0.004, p = 0.013), fusiform gyrus (β = -0.021, 95% CI: -0.037, -0.005, p = 0.009), superior temporal gyrus (β = -0.020, 95% CI: -0.037, -0.002, p = 0.025), temporal pole (β = -0.018, 95% CI: -0.034, -0.003, p = 0.021), and transverse temporal gyrus (β = -0.020, 95% CI: -0.036, -0.004, p = 0.014). After FDR correction, associations for the posterior banks of the superior temporal sulcus, fusiform gyrus, superior temporal gyrus, and transverse temporal gyrus remained significant (FDR p = 0.042 -0.045). Moreover, thinning in the right postcentral gyrus (β = -0.020, 95% CI: -0.036, -0.004, p = 0.012) and right supramarginal gyrus (β = -0.021, 95% CI: -0.038, -0.005, p = 0.013) was significantly linked to higher MBI-C Belief scores, with both remaining significant after FDR correction (FDR p = 0.039). The right insula also exhibited a significant association (β = -0.01736, 95% CI: -0.03374, -0.00112, p = 0.034), retaining significance post-correction (FDR p = 0.037).Table 3 presents the results.

**Table 3.** Detailed Associations Between Cortical Thinning in Specific Brain Subregions and MBI-C Belief Scores.

| Brain Region | B | 95% CI | p | Adj. p |
| --- | --- | --- | --- | --- |
| **Temporal** |  |  |  |  |
| L Entorhinal | -0.017 | -0.032, -0.001 | 0.034 | 0.153 |
| L Fusiform | -0.02 | -0.036, -0.004 | 0.014 | 0.126 |
| R Posterior Banks of Superior Temporal Sulcus | -0.02 | -0.037, -0.004 | 0.013 | **0.042** |
| R Fusiform | -0.021 | -0.037, -0.005 | 0.009 | **0.042** |
| R Superior Temporal | -0.02 | -0.037, -0.002 | 0.025 | **0.045** |
| R Temporal Pole | -0.018 | -0.034, -0.003 | 0.021 | **0.045** |
| R Transverse Temporal | -0.02 | -0.036, -0.004 | 0.014 | **0.042** |
| **Parietal** |  |  |  |  |
| R Postcentral | -0.02 | -0.036, -0.004 | 0.012 | **0.039** |
| R Supramarginal | -0.021 | -0.038, -0.005 | 0.013 | **0.039** |
| **Insula** |  |  |  |  |
| R Insula | -0.01736 | -0.03374, -0.00112 | 0.034 | **0.037** |

*Note.* Models were adjusted for age, education, and global cognition scores. Significant p-values appear bold.
Abbreviations: Adj = Adjusted, L = left, R = right

## Discussion

Our study demonstrates robust cortical thinning across cognitive stages (CN, SCD, and MCI) and a significant association between elevated MBI-C Beliefs scores and atrophy in key brain regions, particularly in the right hemisphere. Pronounced thinning in the temporal lobes—which are crucial for memory and semantic processing—reinforces their vulnerability in early AD (39,40). Additionally, parietal thinning (reflecting impaired spatial awareness and attention) and insular atrophy (linked to disrupted emotional and cognitive integration) underscore the extensive neurodegenerative changes characteristic of AD (29,33,41).

Notably, individuals with SCD exhibited higher MBI-C total and subdomain scores than both CN and MCI participants. This finding may reflect heightened self-awareness or hypervigilance, leading to increased reporting of early NPS that precede overt cognitive impairment (12,42). Elevated symptom reporting may also reflect early affective disturbances and compensatory mechanisms before objective deficits emerge (43).

Consistent with prior reports, our data underscore a strong association between MBI-C Belief scores and cortical thinning in the temporal, parietal, and insular regions. Specifically, bilateral temporal atrophy—among the earliest structural changes in AD—was correlated with higher MBI-C Belief scores, suggesting that loss of neuronal integrity in these regions underlies both cognitive and behavioral disturbances. Additionally, significant associations between right parietal lobe thinning and MBI-C Belief scores highlight the impact on spatial orientation and perception (44), while thinning in the right insula aligns with its role in interoception, emotional regulation, and the genesis of paranoid delusions (45).

Regional analysis in the right hemisphere revealed that cortical thinning in the posterior banks of the superior temporal sulcus, fusiform gyrus, superior temporal gyrus, temporal pole, and transverse temporal gyrus was significantly linked to elevated MBI-C Belief scores. Notably, atrophy in the posterior banks of the superior temporal sulcus—critical for integrating auditory and visual cues in social cognition—may contribute to deficits in social perception and delusional thinking (46). Furthermore, thinning of the temporal pole, integral to emotional regulation and semantic memory, was linked to misinterpretations and delusional behaviors characteristic of MBI (47). Additional significant associations in the right postcentral and supramarginal gyri, regions involved in somatosensory processing and body perception, as well as in the right transverse temporal gyrus, underscore the role of auditory misperceptions in abnormal beliefs (48). Together, these observations point to the right hemisphere’s particular vulnerability in regulating self-perception, sensory integration, and emotional processes—functions often compromised in neurodegenerative disease.

The lateralized pattern of cortical thinning, with more pronounced associations on the right side, aligns with literature suggesting that AD-related pathology can show hemispheric asymmetry in regions governing behavior and emotion (34,49–51). This asymmetry may account for the predominance of NPS, such as apathy and delusions, observed in early dementia. Overall, our findings suggest that MBI may represent both an early behavioral manifestation of cognitive decline and a reflection of underlying structural changes.

## Clinical and Public Health Implications

Our results indicate that the MBI-C could serve as a viable screening tool for identifying subtle neurodegenerative changes. When used alongside established cognitive measures (e.g., MoCA), the MBI-C may enhance diagnostic precision and inform clinical decisions regarding neuroimaging.

Incorporating routine MBI screening into primary care and memory clinics could facilitate earlier detection of behavioral symptoms, supporting timely interventions. Moreover, increasing public awareness of early behavioral alterations—particularly delusions and emotional dysregulation—may reduce stigma and encourage earlier healthcare engagement.

## Limitations

Several limitations warrant consideration. First, although including a Southeast Asian cohort adds diversity, the single-region sample limits generalizability; larger, multi-site, multi-ethnic studies are needed to validate these results. Second, our cross-sectional design prevents definitive causal inferences, highlighting the need for longitudinal studies to delineate the temporal relationships between MBI symptoms and cortical thinning. Third, reliance on self-reported MBI-C data may introduce reporting bias; collecting caregiver input and more objective behavioral data could improve accuracy. Fourth, unmeasured confounders—such as psychiatric conditions, medication use, or vascular risk factors—may have influenced our findings. Finally, focusing solely on cortical thickness narrows the scope of neurodegenerative assessment; future work should integrate additional neuroimaging markers (e.g., white matter integrity, subcortical volumes) to build a more comprehensive picture of the underlying pathophysiology.

## Conclusion

In summary, our study indicates that self-reported MBI, as measured by the MBI-C, is associated with cortical thinning in regions involved in emotional regulation and cognitive processing, particularly within the right hemisphere. These results support the value of behavioral assessments in detecting early signs of neurodegeneration. Longitudinal research in larger, diverse populations will be crucial to clarify how behavioral symptoms and structural changes co-evolve and to optimize early diagnostic and intervention strategies for dementia.

## Funding

This study received funding support from the Strategic Academic Initiative grant from the Lee Kong Chian School of Medicine, Nanyang Technological University, Singapore, National Medical Research Council, Singapore under its Clinician Scientist Award (MOH-CSAINV18nov-0007), Ministry of Education Start-up Grant, Ministry of Education Academic Research Fund Tier 1 (RT02/21) and Ministry of Education Science of Learning Grant (MOESOL2022-0002).

## Data availability statement

Data may be made available upon reasonable request to the corresponding author.

## Ethics approval and consent to participate

Informed consent was obtained from all participants according to the Declaration of Helsinki and local clinical research regulations, and procedures used in the study were in accordance with ethical guidelines. This study has been approved by the NTU Institutional Review Board (NTU-IRB-2021-1036).

## Acknowledgements

The authors express their gratitude to all individuals participating in research at the Dementia Research Centre (Singapore).

## Author Contributions

*Concept and design:* Yi Jin Leow, Nagaendran Kandiah

*Acquisition and interpretation of data:* Yi Jin Leow, Ashwati Vipin, Pricilia Tanoto, Rasyiqah Binte Shaik Mohamed Salim, Bocheng Qiu, Zahinoor Ismail, Nagaendran Kandiah

*Drafting of the manuscript:* Yi Jin Leow, Seyed Ehsan Saffari, Nagaendran Kandiah

*Statistical analysis:* Yi Jin Leow, Seyed Ehsan Saffari

*Supervision:* Nagaendran Kandiah

## Conflict of Interest Disclosures

All authors declare no competing interests.

## Notes

### Competing Interest Statement

The authors have declared no competing interest.

### Clinical Protocols

https://link.springer.com/article/10.14283/jpad.2024.89

### Author Declarations

This study has been approved by the NTU Institutional Review Board (NTU-IRB-2021-1036).

### Summary of Updates

This version includes manuscript following submission to Biological Psychiatry. The title has also been updated for clarity.

